# Association of Habitual Diet Quality and Nutrient Intake with Cognitive Performance in Community-Dwelling Older Adults: A Cross-sectional Study

**DOI:** 10.1101/2025.08.26.25334482

**Authors:** Samitinjaya Dhakal, Nirajan Ghimire, Sophia Bass

## Abstract

**Objectives:** The rapid aging of the U.S. population has raised concerns about age-related cognitive decline and Alzheimer’s disease. As of 2024, 18% of Americans are ≥65 years—up from 12.4% in 2004—contributing to a projected 7.2 million cases of Alzheimer’s disease among older adults in 2025. Diet is a key modifiable factor for cognitive decline. Therefore, we aimed to characterize diet quality and nutrient intake and to examine the associations between specific dietary components and cognitive performance in older adults in the American Midwest.

**Design:** The study was designed as a cross-sectional observational study.

**Setting:** Community-based recruitment in Brookings, South Dakota, and surrounding areas

**Participants:** A final analytical sample of 72 community-dwelling adults aged 65 years and older

**Measurements:** Cognitive performance was assessed using subtests from the Consortium to Establish a Registry for Alzheimer’s Disease (CERAD) battery, evaluating episodic memory (Word List Memory/Recall/Recognition), visuospatial skills (Constructional Praxis), and executive function (Verbal Fluency). A composite cognitive score was calculated from memory and visuospatial subtests. Habitual dietary intake was evaluated using structured 24-hour recalls to calculate nutrient intake and the Healthy Eating Index score, supplemented by the Short HEI questionnaire. Demographics, health history, depressive symptoms (Patient Health Questionnaire-9), and sleep quality (Pittsburgh Sleep Quality Index) were also collected.

**Results:** Participants demonstrated suboptimal diet quality (mean HEI score 54.4 ± 9.4; recommended >80), with only 9.7% meeting fiber recommendations, 11% meeting calcium or vitamin A recommendations, and 1.4% meeting vitamin D requirements. In bivariate comparisons, higher cognitive performance was observed in younger participants (75.5 vs. 79.5 years; p<0.01) and females (78% vs. 50%; p=0.024). Regression models identified significant positive associations between cognitive scores and intakes of dietary fiber (p=0.007), unsaturated fats (mono- and polyunsaturated; p=0.012–0.033), protein (p=0.018), carotenoids (α-carotene, p=0.001; β-carotene, p=0.026; lutein+zeaxanthin, p=0.016), vitamins A (p=0.044) and E (p=0.034), and minerals including magnesium (p=0.006), potassium (p=0.004), copper (p=0.008), zinc (p=0.024), and calcium (p=0.035). Refined grain intake was inversely associated with cognition (p=0.011).

**Conclusion:** In this population, dietary components like fiber and micronutrients were positively associated with better cognitive function, and the overall nutrient intake shortfalls observed highlight the need for targeted dietary interventions to support healthy brain aging.

## 1. Introduction

As of 2024, 18.0% of Americans are 65 or older, up from 12.4% in 2004, far outpacing the 1.4% growth of working-age adults, resulting in a significant rise in overall median age [1]. Aging is the strongest risk factor for Alzheimer’s disease (AD) [2], which affects about 7.2 million Americans aged 65+ in 2025 [3]. In 2025, AD and related dementia care in the U.S. will cost $384 billion, which is more than heart disease or cancer, with Medicare and Medicaid covering two-thirds of the cost [3]. Furthermore, unpaid caregivers provide care worth another $413 billion in addition to the emotional strain [3]. While growing age and genetics are the major contributors [2, 4], emerging evidence suggests many dementia cases may be preventable or, at the very least, delayable [5–7]. The Lancet Commission concluded that up to 40% of global dementia cases are attributable to known, modifiable risk factors, and recent 2024 work extends this estimate to ∼45% when additional factors are included [6, 7].

Among lifestyle factors, dietary modification is one of the most promising and accessible interventions [6–9]. Diet can change key biological pathways associated with AD, with existing evidence showing plant-rich dietary patterns to reduce neuroinflammation and oxidative stress, and processed, high-fat diets to heightened inflammatory signaling and accelerated neurodegeneration [6, 8–12]. Neuroprotective diets rich in in antioxidant and anti-inflammatory compounds are known to combat the neuroinflammation and oxidative stress that drive neurodegeneration (Mediterranean and MIND diets) [9, 13–15] directly, as phytochemicals like polyphenols cross the blood-brain barrier to neutralize reactive oxygen species, and indirectly, as gut microbes ferment dietary fiber into short-chain fatty acids that reduce systemic inflammation [12, 16–19]. However, the cognitive impact of adhering to the Dietary Guidelines for Americans (DGA) [20], which is built on these same principles and is more applicable to the U.S. population, remains largely unexplored, especially in older adults in the American Midwest [21].

To address this gap, the present pilot cross-sectional study investigated habitual dietary intake and cognitive performance in community-dwelling older adults in South Dakota. This study had two primary aims: i) to characterize overall diet quality and nutrient intake in this population by calculating the Healthy Eating Index (HEI) score, and ii) to examine associations of both the HEI score and specific dietary components (including: macronutrients, fiber, refined grains, and key micronutrients such as carotenoids, vitamins, and minerals) with performance across multiple cognitive domains (memory, executive function, and visuospatial ability). This study was exploratory; however, based on prior literature, we anticipated that diets richer in neuroprotective nutrients would correspond with stronger cognitive performance, whereas lower-quality dietary components might show the opposite trend.

## 2. Methods

### 2.1. Study Design and Participants

This study was designed as a cross-sectional observational study to investigate the relationship between habitual dietary patterns and cognitive health. Participants were recruited from October 14, 2024, to March 13, 2025, from the local community in Brookings, South Dakota, and surrounding areas. Eligible individuals were adults aged 65 years and older, able to provide informed consent, and fluent in English. Exclusion criteria included a self-reported diagnosis of a neurodegenerative disease (e.g., Alzheimer’s disease, Parkinson’s disease), a medical condition known to significantly impair cognitive function, or residence in a nursing home or assisted living facility. A total of 76 participants were enrolled in the study. The study was done in accordance with Declaration of Helsinki and was approved by the Institutional Review Board (IRB) at South Dakota State University (IRB-2024-56). All participants provided written informed consent before any study procedures.

### 2.2. Recruitment and Procedures

Recruitment strategy used to reach a broad sample of the older adult population included: distributing flyers at local community centers, libraries, and clinics; posting advertisements on social media platforms; utilizing the Every Door Direct Mail service; and snowball sampling. Interested individuals were screened for eligibility via telephone call. Eligible participants attended a single study visit, lasting approximately 90 minutes at either the South Dakota State University main campus or the Brookings Activity Center. During the visit, the Principal Investigator and a trained researcher administered the cognitive assessment battery [22, 23] and the 24-hour dietary recall interview. All other questionnaires were self-administered by participants using a laptop computer via the REDCap platform [24] as these measures involved sensitive personal information, and self-administration was intended to enhance privacy and comfort for the participants [25].

### 2.3. Cognitive Function Assessment

Cognitive performance was assessed using subtests from the Consortium to Establish a Registry for Alzheimer’s Disease (CERAD) neuropsychological battery [22, 23], a well-validated tool widely used for evaluating episodic memory, visuospatial skills, and executive functions in older adults [22]. These subtests are commonly used in large-scale epidemiological surveys (e.g. NHANES) to explore cognitive functions of older adults. The specific tests that we administered were:

#### 2.3.1. Verbal Fluency (Animal Naming; CERAD J1)

Assesses semantic memory and executive function. Participants were asked to name as many animals as possible in 60 seconds. The score is the total number of unique, correct animals named within the time limit [22, 23, 26].

#### 2.3.2. Word List Memory (CERAD J4)

This task evaluates verbal episodic memory and new learning. Participants were read a list of 10 common, unrelated nouns and immediately asked to recall as many as possible. This process was repeated for a total of three learning trials. The score is the average of all correct words recalled across all three trials (maximum score = 10) [22, 23, 26].

#### 2.3.3. Constructional Praxis (CERAD J5)

This test assesses visuospatial ability and planning. Participants were shown four geometric figures of increasing complexity (circle, diamond, overlapping rectangles, cube), one at a time, and asked to copy each one. Each drawing is scored based on accuracy and the correct representation of all parts, with a maximum possible score of 11 [22, 23, 26].

#### 2.3.4. Word List Recall, Delayed (CERAD J6)

After a delay interval of approximately 5-10 minutes, during which the Constructional Praxis tasks were administered, participants were asked to recall the 10-word list from the earlier memory task. This surprise recall test is a measure of delayed episodic memory, specifically memory consolidation. The score is the total number of words correctly recalled (maximum score = 10) [22, 23, 26].

#### 2.3.5. Word List Recognition (CERAD J7)

Immediately following the delayed recall task, this test assesses delayed recognition memory. Participants were presented with a list of 20 words. 10 original target words and 10 new distractors words and were asked to identify (with a ‘yes’ or ‘no’) whether each word was on the original list. The score is the total number of correct hits (both positive and negative) [22, 23, 26].

#### 2.3.6. Recall of Constructional Praxis, Delayed (CERAD J8)

To assess delayed visuospatial memory, participants were asked, without prior warning, to draw the four geometric figures they had copied earlier. Each recalled figure is scored for accuracy, similar to the immediate copy task [22, 23, 26].

A composite cognitive score was calculated to derive an overall cognitive function score. To create this score, individual scores from each subtest (Word List Memory, Constructional Praxis, Word List Recall, Word List Recognition, and Delayed Recall of Constructional Praxis; J4–J8) were summed. This score was then dichotomized at the sample median to create two groups: Lower Cognitive Performance (LCP) and Higher Cognitive Performance (HCP)

### 2.4. Dietary Assessment

Habitual dietary intake was evaluated using two different methods:

#### 2.4.1. 24-Hour Dietary Recall

The Principal Investigator conducted a structured 24-hour dietary recall interview. Participants were asked to recall all foods and beverages consumed during the preceding day, including portion sizes, preparation methods, and brand names when applicable. Nutrient intake (e.g., calories, macronutrients, micronutrients) was calculated by: first, each reported food and beverage item was disaggregated and assigned a corresponding 8-digit USDA food code and saved as a .csv file. A custom script developed in Python (version 3.12.8) was then utilized to map these food codes to the USDA’s Food and Nutrient Database for Dietary Studies (FNDDS, 2017-2018). The food and nutrient data from the recall were also used to calculate an HEI score for each participant using custom python code [https://github.com/nirajang20/HEI]. The HEI score includes 13 components (9 for adequacy, 4 for moderation) that are summed to produce a total score from 0 to 100, with higher scores indicating better diet quality for that day.

#### 2.4.2. Short Healthy Eating Index (sHEI)

A single day’s intake may not fully represent an individual’s typical diet; therefore, we added the sHEI survey [27]. This validated 23-item questionnaire is a brief instrument to assess the sHEI. This was originally validated with a college population. We selected this tool for its brevity and ease of administration in a community setting. The sHEI score reflects participants’ adherence to the guidelines and provides us with a complementary measure.

### 2.5. Health Questionnaires

#### 2.5.1. Demographics and Health History

A custom questionnaire collected information on age, sex, years of education, race/ethnicity, and self-reported medical history (e.g., hypertension, diabetes, cardiovascular disease) was administered via REDCap.

#### 2.5.2. Patient Health Questionnaire-9 (PHQ-9)

This tool was used to screen for and measure the severity of depressive symptoms. Each item in the tool was scored from 0 (“not at all”) to 3 (“nearly every day”), resulting in a score ranging from 0 to 27. We used this because depressive symptoms are independently associated with both dietary habits and cognitive performance; the total score was then utilized as a covariate in some statistical models to control for its potential confounding effects [28, 29].

#### 2.5.3. Pittsburgh Sleep Quality Index (PSQI)

Subjective sleep quality and disturbances over the past month was assessed using the 19-item self-report questionnaire that evaluates seven components of sleep: subjective quality, latency, duration, efficiency, disturbances, use of sleep medication, and daytime dysfunction. The scores from these components are summed to create a global PSQI score ranging from 0 to 21. Higher scores indicate poorer sleep quality. [30].

### 2.6. Statistical Analysis

All statistical analyses were conducted using R (4.4.1, 2024-06-14 ucrt; R Core Team, 2024). A two-tailed p-value of less than 0.05 was considered the threshold for statistical significance for all tests.

#### 2.6.1. Descriptive Statistics and Group Comparisons

Descriptive statistics were calculated for all demographic, health, cognitive, and dietary variables. Continuous variables were summarized as means and standard deviations (SD), and categorical variables were summarized as frequencies and percentages (%). For bivariate analyses, participants were stratified into two groups (Low cognitive performance, LCP, and High cognitive performance, HCP) based on a median split of their composite cognitive score. To compare characteristics between these two groups, the choice of statistical test was determined by the data type and distribution. For continuous variables, normality was first assessed within each group using the Shapiro-Wilk test. If the data were normally distributed (p > 0.05), group means were compared using an independent samples t-test. If the data were not normally distributed in either group, a Wilcoxon rank-sum test (Mann-Whitney U test) was used. For categorical variables, group differences were assessed using Pearson’s Chi-squared test or Fisher’s exact test.

#### 2.6.2. Multivariate Regression Modeling

A series of generalized linear models with a Gaussian family distribution was used to explore the independent association between specific dietary components and cognitive performance.

Separate models were run for each cognitive outcome (Total Cognitive Score, Memory Composite Score, etc.) as the dependent variable. In each model, a single dietary component (e.g., alpha-carotene, total fat) was entered as the primary independent variable. To control for potential confounding, all models were adjusted for age (as a continuous variable) and sex (as a categorical variable).

## 3. Results

### 3.1. Participant Flow and Sample Characteristics

The recruitment began with an initial phone screening of more than 100 individuals from the local community who expressed interest in the study. From this pool, some were determined to be ineligible and were excluded from participation. The most common reasons for exclusion included: age requirement, residing in a nursing home or assisted living, or declining to participate after the initial screening. This phone screening resulted in a group of 76 eligible individuals, of which 74 participants provided written informed consent and were enrolled. All 74 participants provided written informed consent and were enrolled. Out of 76 participants, 72 participants successfully completed the study visit, including all cognitive assessments and dietary interviews; therefore, the final analytical sample for this study consists of 72 older adults. A detailed visual representation of the participant screening, enrollment, and inclusion in the final analysis is provided in **Figure 1**.

**Figure 1:**
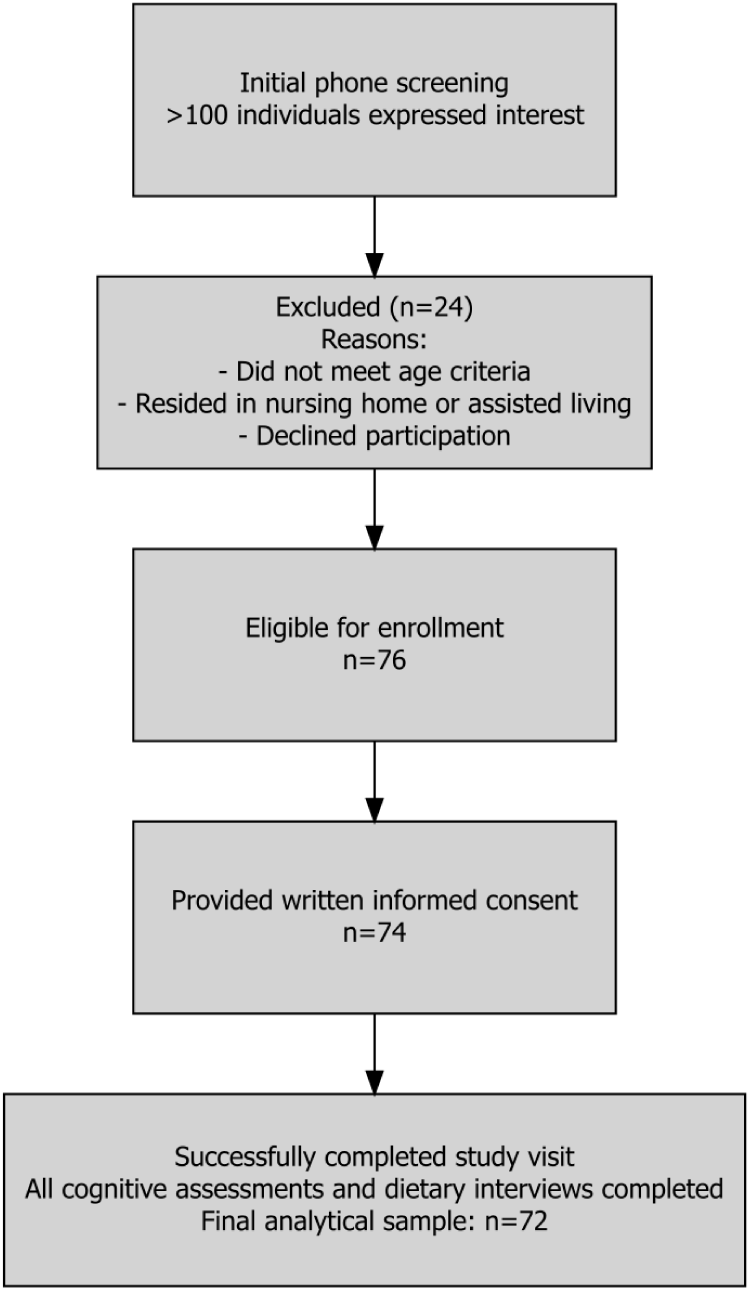
Participant recruitment and enrollment flow diagram.

The demographic, health, and lifestyle characteristics of the final study cohort are summarized in **Table 1**. The average age of the participants was 77.5 years. The sample was predominantly female, with 64% of the cohort. The racial and ethnic composition was largely homogeneous, with all but one of participants identifying as White, non-Hispanic. In terms of self-reported health conditions known to be associated with cognitive function, 49% of participants reported a history of hypertension, 18% reported a diagnosis of diabetes mellitus, and 46% reported hyperlipidemia. The cohort reported good mental health and sleep quality. To perform an initial, unadjusted exploration of factors related to cognitive performance, participants’ characteristics, we stratified participants based on a median split of the composite cognitive score (J4-J8). The comparison table is presented in **Table 1**. The results show that participants in the lower cognitive performance group were significantly older than those in the higher group (mean age = 79.5 ± 6.6 in LCP vs. 75.5 ± 4.9 years in HCP; p < 0.01). A significant difference was also observed in the distribution of sex between the groups (p=0.024), with a higher proportion of females in the high-performance group (78%) compared to the low-performance group (50%). No statistically significant differences were observed between the groups for family history of dementia, alcohol use, or the prevalence of individual health conditions (**Table 1**).

**Table 1:**
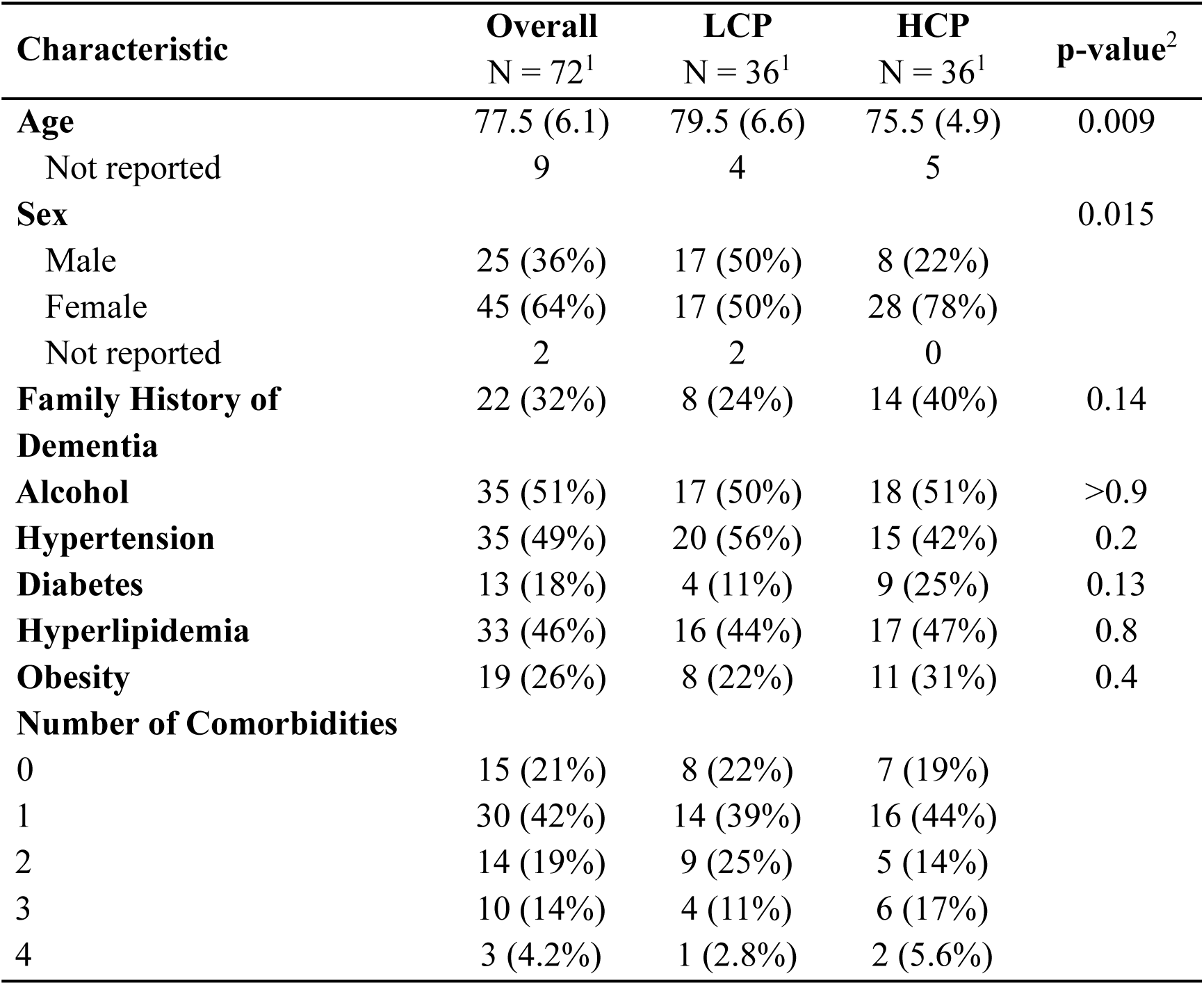
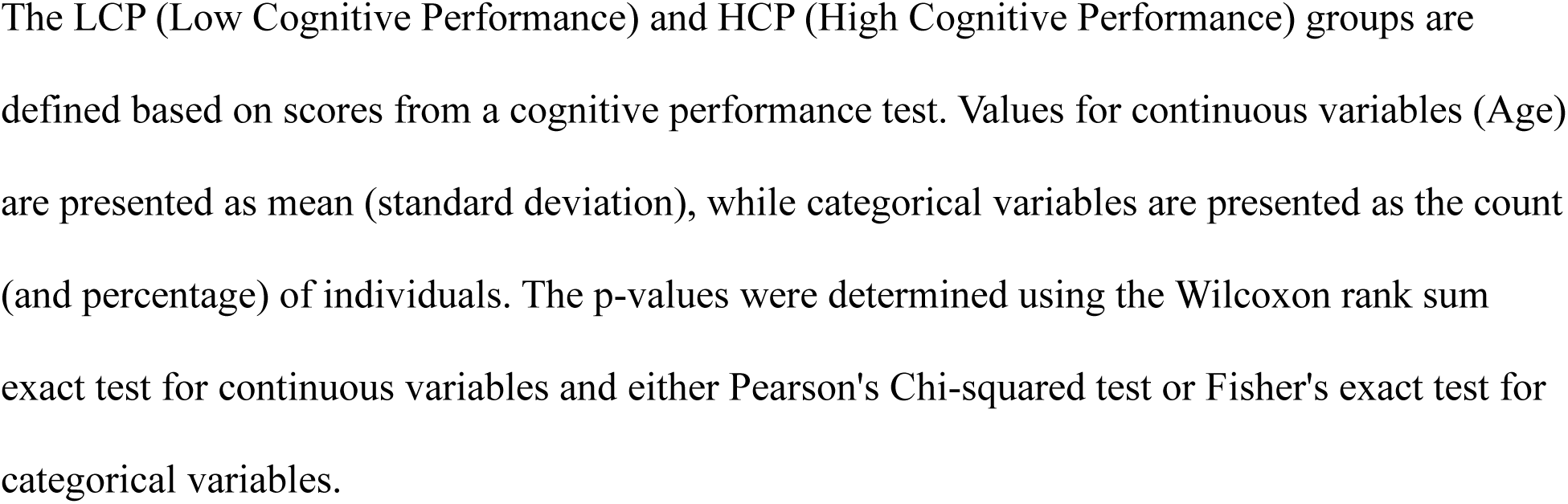
Participant Characteristics by Cognitive Performance Group.

### 3.2. Cognitive Performance Outcomes

The cognitive performance scores for the overall cohort and stratified by cognitive group are presented in **Table 2**. As expected, the group designated with HCP demonstrated significantly better scores across all assessed domains compared to the LCP group. The mean Total Cognitive Score, a composite of all tests excluding verbal fluency, was substantially higher in the HCP (52.0 ± 4.0) compared to the LCP (38.1 ± 5.5; p < 0.001). This pattern was consistent across the primary cognitive domains. The HCP showed significantly higher scores on the Memory Composite Score (31.8 ± 3.7 vs. 22.6 ± 4.0; p < 0.001) and the Visuospatial Composite Score (20.2 ± 1.8 vs. 15.4 ± 3.7; p < 0.001). Analysis of individual subtests showed that the differences were driven by superior performance in immediate word list memory, constructional praxis copy, delayed word recall, word recognition, and delayed praxis recall (all, p < 0.001). The difference in the Executive Functioning Score, measured by verbal fluency, was also higher and close to significance (22.0 ± 7.4 vs. 18.6 ± 5.3; p = 0.056).

**Table 2:** Comparison of Cognitive Outcomes Between Cognitive Performance Groups.

| Cognitive Outcomes | Overall<br>N = 72<sup>1</sup> | LCP<br>N = 36<sup>1</sup> | HCP<br>N = 36<sup>1</sup> | p-value<sup>2</sup> |
| --- | --- | --- | --- | --- |
| Word List Memory (Immediate) | 5.2 (2.8) | 3.4 (2.2) | 7.1 (2.0) | <0.001 |

| **Cognitive Outcomes** | **Overall**<br>N = 72<sup>1</sup> | **LCP**<br>N = 36<sup>1</sup> | **HCP**<br>N = 36<sup>1</sup> | **p-value**<sup>2</sup> |
| --- | --- | --- | --- | --- |
| **Constructional Praxis (Copy)** | 9.7 (1.7) | 8.9 (1.9) | 10.5 (0.8) | <0.001 |
| **Word List Recall (Delayed)** | 3.9 (2.6) | 2.0 (1.9) | 5.8 (1.4) | <0.001 |
| **Word List Recognition** | 18.1 (1.7) | 17.3 (2.0) | 18.9 (1.0) | <0.001 |
| **Constructional Praxis (Delayed Recall)** | 8.1 (2.5) | 6.6 (2.3) | 9.7 (1.4) | <0.001 |
| **Total Cognitive Score** | 45.0 (8.5) | 38.1 (5.5) | 52.0 (4.0) | <0.001 |
| **Memory Composite Score** | 27.2 (6.0) | 22.6 (4.0) | 31.8 (3.7) | <0.001 |
| **Visuospatial Composite Score** | 17.8 (3.7) | 15.4 (3.7) | 20.2 (1.8) | <0.001 |
| **Executive Functioning Score** | 20.3 (6.6) | 18.6 (5.3) | 22.0 (7.4) | 0.056 |

Values are presented as mean (standard deviation). The p-values were determined using the
Wilcoxon rank sum test. LCP: Low Cognitive Performance; HCP: High Cognitive Performance

### 3.3. Macronutrients and Energy

The dietary intake, diet quality scores, and adherence to the dietary recommendations are detailed in **Table 3**. In unadjusted analyses comparing the HCP and LCP groups, no statistically significant differences were observed for any measure of macronutrients, energy, or quality. However, after adjusting for age and sex in regression models, several of these components showed up as significant predictors of the Total Cognitive Score. Higher intakes of total fat (p=0.010), saturated fat (p=0.026), monounsaturated fat (p=0.012), polyunsaturated fat (p=0.033), protein (p=0.018), dietary fiber (p=0.007), and total energy (p=0.015) were all significantly associated with better cognitive performance. This pattern of positive associations was also present for the Memory Composite Score. Furthermore, higher intake of monounsaturated fat was also significantly associated with better Executive Functioning Scores (p=0.027). Conversely, a higher intake of refined grains was significantly associated with a lower cognitive score (p=0.011). The average daily energy intake for the cohort was 1662.8kcal. The HCP reported a slightly higher intake of energy (1,796.1 vs. 1,529.5 kcal), dietary fiber (18.0 vs. 14.0 g/day), and total fat (81.5 vs. 64.9 g/day) compared to the LCP. Overall diet quality, as measured by the HEI-2015, was nearly identical between the groups (54.8 vs. 54.0, p=0.5), with a mean score for the entire cohort of 54.4, indicating a need for a major improvement. An analysis of adherence to specific dietary recommendations also showed low compliance across the study population. A majority (85%) met the Acceptable Macronutrient Distribution Range for protein, but less than half (49%) met the range for carbohydrates, and only 32% met the range for fat. Adherence was similarly low for the recommendation to limit saturated fat intake to less than 10% of total calories, with only 31% of participants meeting this target. The most notable finding was the extremely low adherence to the daily fiber recommendation, which was met by only 9.7% of the cohort. There were no significant differences in the proportion of individuals meeting any of these guidelines between the two cognitive performance groups (all p > 0.2).

**Table 3:** Comparison of Dietary Intake and Quality Between Cognitive Performance Groups.

| **Dietary Variable** |  | **Overall**<br>N = 72<sup>1</sup> | **LCP**<br>N = 36<sup>1</sup> | **HCP**<br>N = 36<sup>1</sup> | **p-value**<sup>2</sup> |
| --- | --- | --- | --- | --- | --- |
| **Dietary Intake & Quality** | Energy (kcal/day) | 1,662.8<br>(978.0) | 1,529.5<br>(824.5) | 1,796.1<br>(1,106.2) | 0.3 |
|  | Protein (g/day) | 61.3 (40.0) | 57.1 (32.5) | 65.6 (46.5) | 0.6 |
|  | Carbohydrate (g/day) | 192.0 (128.1) | 176.0 (85.5) | 207.9 (159.6) | 0.7 |
|  | Total Sugars (g/day) | 84.8 (72.5) | 76.5 (48.2) | 93.0 (90.6) | 0.6 |
|  | Dietary Fiber (g/day) | 16.0 (9.7) | 14.0 (7.7) | 18.0 (11.1) | 0.2 |
|  | Total Fat (g/day) | 73.2 (53.8) | 64.9 (43.1) | 81.5 (62.3) | 0.2 |
|  | Saturated Fat (g/day) | 22.4 (16.1) | 19.9 (13.2) | 24.9 (18.4) | 0.2 |
|  | Monounsaturated Fat (g/day) | 26.6 (24.1) | 23.4 (18.6) | 29.8 (28.4) | 0.2 |
|  | Polyunsaturated Fat (g/day) | 18.1 (14.5) | 16.0 (11.2) | 20.2 (17.0) | 0.4 |
| **Guideline Adherence (% Meeting)** | HEI Total Score | 54.4 (9.4) | 54.0 (9.3) | 54.8 (9.6) | 0.5 |
|  | sHEI Total Score | 52.1 (9.7) | 53.0 (9.0) | 51.1 (10.5) | 0.4 |
|  | Protein AMDR (10-35%) | 61 (85%) | 32 (89%) | 29 (81%) | 0.3 |
|  | Carbohydrate AMDR (45-65%) | 35 (49%) | 20 (56%) | 15 (42%) | 0.2 |
|  | Fat AMDR (20-35%) | 23 (32%) | 14 (39%) | 9 (25%) | 0.2 |
|  | Saturated Fat (<10% kcal) | 22 (31%) | 13 (36%) | 9 (25%) | 0.3 |

| **Dietary Variable** | **Overall**<br>N = 72<sup>1</sup> | **LCP**<br>N = 36<sup>1</sup> | **HCP**<br>N = 36<sup>1</sup> | **p-value**<sup>2</sup> |
| --- | --- | --- | --- | --- |
| Fiber Recommendation | 7 (9.7%) | 3 (8.3%) | 4 (11%) | >0.9 |

Values for dietary intake and quality variables are presented as mean (standard deviation), unless otherwise noted. Values for guideline adherence are presented as the count (and percentage) of individuals meeting the respective guideline. The p-values were determined using the Wilcoxon rank sum exact test or the Wilcoxon rank sum test. LCP: Low Cognitive Performance; HCP: High Cognitive Performance; AMDR: Acceptable Macronutrient Distribution Range; HEI: Healthy Eating Index; sHEI: Short Healthy Eating Index

### 3.4. Micronutrient intake

A detailed comparison of micronutrient intakes between the cognitive performance groups is presented in **Table 4**. Most individuals’ nutrient intakes did not differ significantly between the groups in unadjusted tests, but a clear and consistent trend was observed. The HCP group had numerically higher mean intakes for the majority of vitamins and minerals analyzed. This trend reached statistical significance for several key carotenoids. Specifically, the high-performance group consumed significantly more alpha-carotene (932.6 vs. 199.2 mcg, p=0.004), beta-carotene (3,853.2 vs. 1,424.1 mcg, *p*=0.012), and lutein + zeaxanthin (2,259.7 vs. 1,597.8 mcg, p=0.016) compared to the low-performance group. To further investigate these associations while controlling for age and sex, we conducted a series of generalized linear models. The regression analyses confirmed the observed trend. Higher intake of several micronutrients was significantly associated with a higher Total Cognitive score. These included alpha-carotene (p=0.001), beta-carotene (p=0.026), cryptoxanthin (p=0.032), Vitamin A (p=0.044), and Vitamin E (p=0.034), phosphorus (p=0.001), potassium (p=0.004), copper (p=0.008), magnesium (p=0.006), zinc (p=0.024), calcium (p=0.035), and sodium (p=0.035). Key B-vitamins: riboflavin (p=0.015), folate from food (p=0.019), and choline (p=0.006), were also positively associated with cognitive scores. This pattern of positive associations was also present for the Memory Composite Score, where higher intakes of alpha-carotene, phosphorus, sodium, Vitamin E, copper, choline, riboflavin, folate, zinc, beta-carotene, and calcium were all significantly associated with better scores. For the Visuospatial Composite Score, higher intakes of potassium and choline were significantly associated with better performance.

**Table 4.** Comparison of Daily Micronutrient Intake Between Cognitive Performance Groups.

| **Micronutrient** | **LCP** | **HCP** | ***p*-value** |
| --- | --- | --- | --- |
| **<u>Vitamins</u>** |  |  |  |
| Vitamin A (RAE, mcg) | 474.0 (346.5) | 713.3 (628.5) | 0.114 |
| Vitamin C (mg) | 83.2 (104.2) | 79.0 (78.1) | 0.750 |
| Vitamin D (mcg) | 3.7 (2.9) | 4.5 (5.2) | 0.906 |
| Vitamin E (total, mg) | 7.3 (4.7) | 9.9 (9.6) | 0.338 |
| Vitamin E (added, mg) | 0.8 (2.9) | 0.7 (2.8) | 0.734 |
| Vitamin K (mcg) | 104.7 (188.2) | 127.7 (148.0) | 0.060 |
| Thiamin (mg) | 1.3 (0.7) | 1.4 (1.0) | 0.924 |
| Riboflavin (mg) | 1.5 (0.8) | 1.8 (1.1) | 0.289 |
| Niacin (mg) | 18.6 (11.5) | 20.1 (17.5) | 0.666 |
| Vitamin B6 (mg) | 1.4 (0.8) | 1.5 (0.9) | 0.767 |
| Folate (total, mcg) | 295.8 (177.0) | 319.9 (191.8) | 0.618 |
| Folate (from food, mcg) | 167.5 (101.5) | 207.8 (130.4) | 0.123 |
| Folic Acid (mcg) | 128.4 (136.6) | 112.0 (107.5) | 0.897 |
| Vitamin B12 (total, mcg) | 2.9 (2.3) | 2.9 (2.7) | 0.853 |
| Vitamin B12 (added, mcg) | 0.5 (1.0) | 0.3 (0.7) | 0.252 |
| Choline (mg) | 239.4 (148.0) | 264.0 (195.3) | 0.793 |
| **<u>Carotenoids</u>** |  |  |  |
| α-Carotene (mcg) | 199.2 (437.9) | 932.6 (1276.2) | **0.004** |
| β-Carotene (mcg) | 1424.1 (2247.8) | 3853.2 (4539.2) | **0.012** |
| β-Cryptoxanthin (mcg) | 76.4 (150.2) | 233.0 (556.6) | 0.577 |
| Lutein + Zeaxanthin (mcg) | 1597.8 (4166.2) | 2259.7 (4134.9) | **0.016** |
| Lycopene (mcg) | 2068.8 (5133.2) | 1979.6 (3657.6) | 0.527 |
| Retinol (mcg) | 344.1 (282.8) | 344.0 (323.8) | 0.826 |
| **<u>Minerals</u>** |  |  |  |
| Calcium (mg) | 629.7 (456.3) | 789.0 (582.0) | 0.195 |
| Copper (mg) | 0.9 (0.5) | 1.1 (0.7) | 0.289 |
| Iron (mg) | 10.3 (6.2) | 10.9 (7.1) | 0.784 |
| Magnesium (mg) | 226.1 (142.9) | 275.9 (185.9) | 0.128 |
| Phosphorus (mg) | 948.2 (539.1) | 1124.9 (675.6) | 0.158 |
| Potassium (mg) | 1922.6 (1039.8) | 2345.7 (1313.9) | 0.133 |
| Selenium (mcg) | 79.6 (44.0) | 87.0 (66.9) | 0.978 |
| Zinc (mg) | 7.8 (5.4) | 8.5 (6.2) | 0.436 |

Values are mean (standard deviation). The *p*-values were determined using the Wilcoxon rank sum exact test or the Wilcoxon rank sum test. Bolded *p*-values indicate statistical significance
(*p*<0.05). LCP: Low Cognitive Performance; HCP: High Cognitive Performance; RAE: Retinol
Activity Equivalents; mcg: micrograms; mg: milligrams

However, it is important to highlight that a substantial portion of the cohort was not meeting nutritional recommendations. Intake was generally sufficient for phosphorus (65% meeting RDA) and selenium (67% meeting RDA), but adequacy was low for many other critical nutrients. Only 11% of participants met the recommendation for Vitamin A, 11% for Vitamin E, and only 1.4% met the RDA for Vitamin D. Similarly, adherence was low for essential minerals like calcium (11% meeting RDA) and potassium (9.7% meeting RDA). There were no statistically significant differences in the proportion of individuals meeting these recommendations between the two cognitive performance groups.

## 4. Discussion

The increasing burden of cognitive diseases represents a major public health challenge that is driven by the aging population and limited prevention options [2, 31]. Existing evidence suggests that modifiable lifestyle factors, including diet, may offer an important way to prevent or delay cognitive decline[5–7, 9, 10, 31]. The present pilot study explored habitual dietary intake and cognitive performance in the community-dwelling older adults in South Dakota. The principal findings from this pilot study show that the adherence to the dietary guidelines was overall critically low among the overall cohort but higher intakes of specific macronutrients, such as protein, fiber, and healthy fats; and key micronutrients, such as: carotenoids, vitamins A and E, choline, magnesium, and potassium were significantly associated with better cognitive function after adjusting for age and sex. Conversely, a higher intake of refined grains was associated with poorer cognitive performance.

The demographic profile of our cohort is broadly consistent with other community-based studies of older adults in the U.S [32–34]. Our population was older, with an average age of 77.5 years, and was predominantly female, and reflected the demographic landscape of the recruitment area. Chronic health conditions were common, with nearly half reporting hypertension or hyperlipidemia and almost one-fifth reporting diabetes, aligning with national prevalence estimates in older adults [35]. These comorbidities are important to discuss because of their well-established associations with cognitive impairment through pathways of vascular dysfunction, neuroinflammation, and metabolic dysregulation [36–38]. Interestingly, our cohort reported minimal depressive symptoms and relatively good sleep quality, which minimizes the potential for confounding as both of these factors are independently linked to both dietary patterns and cognitive outcomes [39–41].

Participants with higher composite cognitive scores showed superior performance across nearly all domains, including memory, visuospatial ability, and recognition tasks. The largest group differences were present in episodic memory and visuospatial tasks, which are domains particularly vulnerable to age-related decline and early Alzheimer’s pathology [42, 43]. In contrast, the between-group difference in executive function, measured by verbal fluency, did not reach traditional statistical significance. One previous study reported that semantic fluency tends to be more resilient to age-related decline relative to memory-based measures [44]. Rönnlund et al 2005 analyzed both cross-sectional and longitudinal data from a large, population-based sample of adults. They saw that semantic memory (recalling facts and general knowledge actually improved through midlife and remained largely stable, with significant declines only apparent after very old age.

We observed a handful of associations between the nutrients and cognitive performance after adjustment for age and sex. The most notable ones were the positive association with dietary fiber intake and the negative association with refined grains. This is biologically plausible. Dietary fiber is fermented by the gut microbiota to produce short-chain fatty acids like butyrate, and other metabolites which are known to cross the blood-brain barrier, reduce neuroinflammation, and enhance synaptic plasticity [45]. The inverse relationship with refined grains, which are low in fiber and high in glycemic load, supports the same conclusion as well. Mechanistically, consumption of refined grains can lead to post-prandial hyperglycemia/insulinemia, oxidative stress, and systemic inflammation, which is linked to neurodegenerative diseases [46, 47].

Furthermore, carotenoids such α-carotene, β-carotene, and lutein/zeaxanthin were some predictors of cognitive outcomes. Studies have shown previously that carotenoids accumulate in neural tissue, where they function as potent antioxidants and can modulate cell signaling [48, 49]. A cohort study by Johnson et al. 2013, analyzed post-mortem tissue from the Georgia Centenarian Study. They found that lutein and zeaxanthin were preferentially concentrated in the brain compared to other carotenoids. In the same cohort, higher concentrations of brain lutein were associated with better scores across a range of premortem cognitive tests. These nutrients protect neurons from oxidative stress and inflammation [48]. This can also be a contributing factor of the MIND and Mediterranean diets as they are rich in different plants [13–15].

Similarly, higher intakes of vitamin A and E, were linked with better cognition in our cohort. This was seen in previous meta-analyses that dietary vitamin E is protective against age-related cognitive decline [50], however, supplementation trials have produced mixed results [51].

Beyond antioxidants, several minerals showed significant associations, like magnesium, potassium, zinc, and copper. They all positively correlated with cognitive performance overall. This is also biologically plausible because of their roles in neurotransmission, synaptic plasticity, and vascular function [52–54]. Magnesium deficiency, for example, has been linked to impaired learning and memory, while adequate potassium intake may support vascular health for cerebral perfusion[52, 53]. Similarly, B-vitamins and choline (also an acetylcholine precursor), associated with the one-carbon metabolism and homocysteine regulation, were associated with better cognitive outcomes [54].

While we saw interesting and positive nutrient and cognition associations, overall diet quality in this cohort was suboptimal, with a mean HEI score well below the recommended target and widespread nutrient shortfalls [21]. Very few participants met important micronutrient and fiber recommendations. These gaps are concerning, given consistent evidence linking inadequate intake of these nutrients to accelerated cognitive decline [50]. The fact that our participants were relatively healthy and community-dwelling and still demonstrated such poor adherence shows the challenge of achieving nutrient adequacy in late life and highlights opportunities for targeted dietary interventions. This study had some limitations. First, the cross-sectional design precludes causal inferences and, therefore, there is a possibility of reverse causation. Second, the modest sample size and ethnically homogenous composition of the sample limits the generalizability of our findings to other populations. Finally, although we used multiple dietary assessment techniques to mitigate biases, the diet data are self-reported, which are susceptible to measurement error and recall biases. Acknowledging these limitations, our study has yielded several important findings. Our study suggests that nutrition as a potential modifiable factor in cognitive aging, with diets richer in fiber, carotenoids, unsaturated fats, and lean protein with lower amounts of refined grains may support better cognitive outcomes. The nutrient inadequacy in the consumption by our cohort highlights the need for targeted dietary counseling and interventions to help older adults meet recommendations for micronutrients and fiber. Future longitudinal and interventional studies are warranted to test whether improving diet quality can slow cognitive decline and promote healthy brain aging.

## Data Availability

All data produced in the present study are available upon reasonable request to the authors

## Abbreviations

AD: Alzheimer’s Disease
AMDR: Acceptable Macronutrient Distribution Range
CERAD: Consortium to Establish a Registry for Alzheimer’s Disease
HCP: High Cognitive Performance
IRB: Institutional Review Board
LCP: Low Cognitive Performance
PHQ-9: Patient Health Questionnaire-9
PSQI: Pittsburgh Sleep Quality Index
RDA: Recommended Dietary Allowance
sHEI: Short Healthy Eating Index

## Acknowledgements

The authors would like to express their sincere gratitude to the participants of this study for their time and valuable contributions. We would also like to thank the Brookings Activity Center for allowing us to use their facilities for surveys

## Authorship contribution statement

### Samitinjaya Dhakal

Conceptualization, Funding acquisition, Methodology, Supervision, Project administration, Resources, Software, Data curation, Formal analysis, Writing – original draft, Writing – review & editing

### Nirajan Ghimire

Investigation, Data curation, Software, Formal analysis

### Sophia Bass

Investigation, Data curation

## Declaration of Generative AI and AI-assisted technologies in the writing process

During the preparation of this work, the authors used LLM to assist with sentence structure and grammar. All content was subsequently reviewed and edited by the authors, who take full responsibility for the final version of the manuscript.

